# Correspondence between morphological similarity of the left lateral orbitofrontal cortex and neurotransmitter systems in adolescent males with autism

**DOI:** 10.64898/2025.12.17.25342383

**Authors:** Huashuang Zhang, Junle Li, Chensheng Hou, Yilin Huang, Lisha Ma, Bincan Xiong, Jinhui Wang, Xuchu Weng

## Abstract

**Background:** Autism spectrum disorder (ASD) is a neurodevelopmental condition marked by pronounced heterogeneity in brain structure, which limits the development of targeted interventions. Morphological brain networks (MBNs) enable mapping of coordinated structural features across brain regions at the individual level. However, the specific organization of such networks in ASD and their potential relationships with underlying neurotransmitter systems remain largely unexplored.

**Aims:** To characterize alterations in cortical thickness-based MBNs among adolescent males with ASD and to test whether these network changes spatially correspond to normative PET-derived neurotransmitter receptor/transporter maps.

**Methods:** In this case-control study, T1-weighted MRI data from 424 adolescent males (207 ASD, 217 typically developing) in the Autism Brain Imaging Data Exchange were analyzed. MBNs were constructed using interregional cortical thickness similarity quantified by Jensen–Shannon divergence. Graph-theoretical metrics were computed and group differences were assessed with permutation tests controlling for age and IQ. Spatial correlations between the left lateral orbitofrontal morphological similarity and atlas-based neurotransmitter maps were investigated using the JuSpace toolbox.

**Results:** Adolescent males with ASD showed increased normalized clustering coefficient (t=2.40, p=0.020, FDR corrected) and reduced nodal measures in the left lateral orbitofrontal cortex (OFC) including degree centrality, PageRank centrality, and betweenness centrality (all p<0.001, FDR corrected). Morphological similarity analysis revealed decreased OFC-based similarity with 65 brain regions. Furthermore, the OFC-related morphological similarity was associated with the spatial distributions of neurotransmitter systems, with GABAa, 5-HT1a, and μ-opioid receptors remaining significant after FDR correction.

**Conclusions:** These findings highlight the left lateral OFC as a structural key hub in adolescent males with ASD, linking left lateral OFC-based morphological similarity to neurotransmitter systems and providing a potential neurobiological basis for targeted interventions in this population.

**What is already known on this topic:** Structural brain alterations and heterogeneity in autism spectrum disorder (ASD) are well established, but little is known about how individual brain networks relate to neurochemical systems that may guide treatment targets.

**What this study adds:** This study adds to existing knowledge by identifying the left lateral orbitofrontal cortex (OFC) as a disrupted structural hub in adolescent males with ASD and demonstrating its morphological similarity alterations are linked to GABAa, 5-HT1a, and μ-opioid receptor systems.

**How this study might affect research, practice or policy:** By establishing spatial correspondence between left lateral OFC morphological similarity and GABAa, 5-HT1a, and μ-opioid receptor systems, this work may provide potential neurobiological markers and targets for circuit-based interventions in adolescent males with ASD.

## Introduction

Autism spectrum disorder (ASD) is a heterogeneous neurodevelopmental condition, characterized by social-communication impairments and restricted, repetitive behaviors.^1^ Marked heterogeneity across clinical presentation, cognitive profiles, neuroimaging patterns, and molecular mechanisms complicates the search for robust, individualized biomarkers and effective interventions.^2^

In response, recent research has shifted from group-averaged models toward individualized brain analyses.^3^ Among these, morphological brain networks (MBNs) provide a promising framework for mapping coordinated anatomical features across regions within individuals, thereby capturing idiosyncratic structural organization often obscured by traditional methods. ^45^ Unlike conventional morphometric covariance networks that rely on across-subject correlations, MBNs quantify intra-subject regional similarity using metrics such as Jensen–Shannon divergence, enhancing sensitivity to individual-level variation.^6^

Although a limited number of studies have applied MBNs in ASD, most using gray-matter volume.^7–10^ In contrast, we employed cortical thickness, which is more developmentally sensitive and biologically interpretable.^11^ Cortical thickness in ASD shows atypical trajectories—early overgrowth followed by premature thinning in adolescence—and relates to core symptom dimensions, and maps onto genes enriched for synaptic transmission, underscoring its utility for characterizing structural heterogeneity.^12^

Adolescence is a critical window marked by synaptic pruning, dendritic remodeling, and intracortical myelination that reshape prefrontal circuits; sex- and age-specific effects further amplify variability, making adolescent males a well-suited subgroup for reducing confounding factors.^13^

Beyond macroscopic topology, recent studies highlight the value of linking structural imaging with molecular systems.^13–16^ Normative positron emission tomography (PET)-derived neurotransmitter maps provide spatial templates for testing whether ASD-related network alterations align with underlying neurotransmitter architectures, enabling cross-scale evaluation of structural–neurochemical correspondence.^15^

To our knowledge, this is the first study to apply cortical-thickness-based MBNs to adolescent males with ASD to (i) identify alterations in network topology and (ii) test their spatial correspondence with normative PET neurotransmitter maps. This framework aimed to bridge structural differences with neurochemical mechanisms and may inform individualized, circuit-based interventions among this population.

## Methods

### Participants

In this case–control study, the participants were obtained from Autism Brain Imaging Data Exchange (ABIDE) datasets (http://fcon_1000.projects.nitrc.org/indi/abide/). Detailed descriptions of evaluation protocols, phenotypic data, data provenance, scanner acquisition parameters and Ethics Committee are provided in this public dataset.^17^ The imaging sample selection in the current study was based on a previous study.^17^ Inclusion criteria were the same as the previous work of Di Martino et al.^17^: (1) age range from 12-18 years old; (2) due to the small percentage of females (<10%) in the datasets, only male subjects were included; (3) only sites where at least 75% subjects have a full scale Intelligence quotient (FSIQ) score were included; (4) only subjects with FSIQ within mean C±C 2SD (108C±C15) were included; (5) each subject had an anatomical image; (6) Sites with fewer than 10 subjects in total were excluded. After these criteria were applied, a total of 424 subjects (207 ASD and 217 TD) recruited from 13 sites were included in the final analysis. We selected the 12–18-year window to optimize sample size and control for potential age effects.^18^ Participant demographics are provided in **Table 1**. The detailed information about the process of participant selection was shown in supplementary materials (**Figure S1**).

**Table 1.** The demographic and psychiatric measurements of included participants.

| | TD (n=217) | ASD (n=207) | $t/\chi^2$ | p-value | Effect size |
| --- | --- | --- | --- | --- | --- |
| Age (years) | 14.68 ± 1.73 | 14.64 ± 1.77 | $t=0.235$ | 0.814 | 0.023 |
| Handedness<br>(right/left/mixed/N/A) | 158/13/5/41 | 153/16/5/33 | $\chi^2=1.020$ | 0.800 | 0.049 |
| Image quality scores (3/4/5) | 69/72/76 | 64/75/68 | $\chi^2=0.458$ | 0.795 | 0.033 |
| IQ |  |  |  |  |  |
| Full scale IQ | 109.64 ± 12.33 | 105.54 ± 14.07 | t=3.185 | 0.002 | 0.310 |
| Performance IQ | 106.64 ± 12.93 | 105.29 ± 13.93 | t=1.033 | 0.302 | 0.101 |
| Verbal IQ | 111.65 ± 13.39 | 106.56 ± 16.04 | t=3.539 | 0.000 | 0.345 |
| ADOS (Total Score) | - | 10.00 ± 12.25 | | | |

Note: Only 98 ASD patients were included in the ADOS total score analysis due to missing scores of some patients. ASD: Autism spectrum disorder; TD: Typically developing; ADOS: Autism Diagnostic Observation Schedule; IQ: Intelligence quotient.

### Image pre-processing

All T1-weighted images underwent a standard preprocessing pipeline using the CAT12 toolbox (http://dbm.neuro.uni-jena.de/cat12/) based on the SPM12 software (https://www.fil.ion.ucl.ac.uk/spm/software/spm12/). Firstly, images from both ASD and TD groups were segmented into gray matter (GM), white matter (WM) and cerebrospinal fluid (CSF) using customized pediatric tissue probability maps generated with the TOM8 toolbox (https://neuro-jena.github.io/software.html#tom). The segmentation incorporated a Hidden Markov random field model to account for partial volume effects.^19^ Subsequently, a map of the distance from WM to the inner boundary of GM was computed,^20^ enabling estimation of GM thickness at the outer boundary.

This thickness was then projected back to the inner boundary to generate vertex-wise cortical thickness maps. The central (mid-thickness) surface was reconstructed at 50% between GM/WM and pial boundaries. Topological defects were corrected using a spherical harmonics-based approach,^21^ and the surface was mapped onto a standard spherical coordinate system for inter-subject alignment.^22^ Surface registration was performed using CAT12’s default surface-based registration approach.^23^ Cortical thickness maps were finally resampled onto the fsaverage template and smoothed using a 12-mm FWHM Gaussian kernel.

### Quality control (QC)

To ensure the integrity and consistency of morphometric data across multiple sites, we conducted a QC process incorporating both automated and manual procedures. Automated QC relied on outputs from the CAT12 pipeline, including surface reconstruction errors, topology flags, and other image quality indices. Scans with failed processing or poor-quality indices were excluded. In addition, two independent reviewers conducted visual inspections to identify motion artifacts, surface-surface intersections, and inaccuracies in GM/WM boundary placement. Only scans judged acceptable by both reviewers were included in the final dataset. Group-level comparisons of image quality indices indicated no systematic differences between ASD and TD participants (**Table 1**).

### Construction of single-subject MBNs

We constructed MBNs based on cortical thickness for each participant in the present study (**Figure 1**).

**Figure 1.**
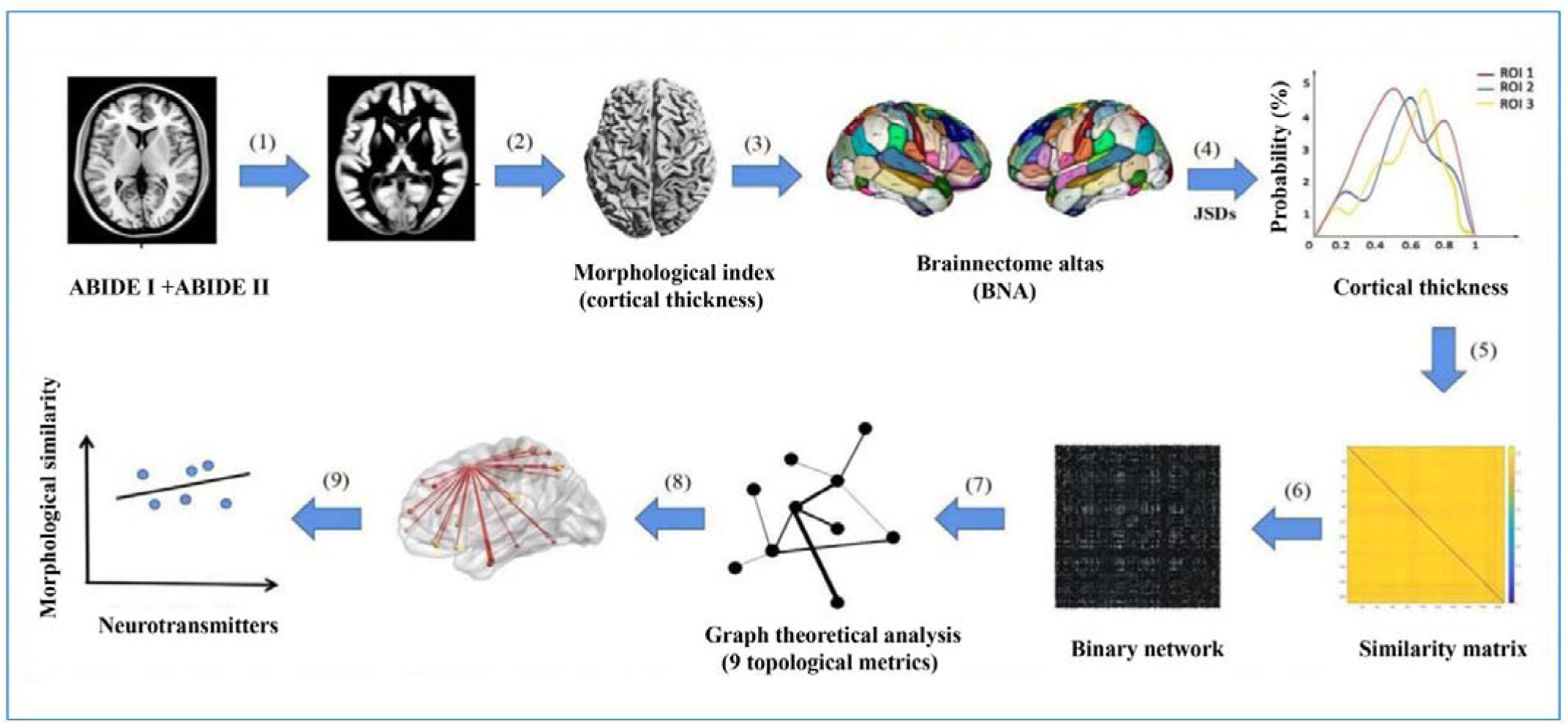
Overview of analysis pipeline. (1) Individual T1-weighted images from ABIDE I and II were segmented into gray matter, white matter, and cerebrospinal fluid; (2) cortical thickness was estimated; (3) the cortex was parcellated into regions according to the Brainnetome Atlas; (4) cortical thickness values within each region were extracted to estimate their probability distributions; (5) pairwise morphological similarity between regions was computed using JSDs, yielding a similarity matrix; (6) the similarity matrix was thresholded to generate binary networks; (7) graph-theoretical measures were calculated to characterize global and nodal topology; (8) based on altered nodal properties in the left lateral orbitofrontal cortex (OFC), we further examined its morphological similarity with other regions; (9) correlations were performed between morphological similarity and neurotransmitter distributions. Autism Brain Imaging Data Exchange; JSDs, Jensen-Shannon divergence similarity; OFC, orbitofrontal cortex.

### Definition of network nodes

To construct MBN for each participant, the brain was segmented into 210 cortical regions of interest (ROIs) using the Brainnetome atlas.^24^ Each ROI was defined as a node of the network. Consequently, a regional similarity matrix was constructed for each participant.

### Definition of network edges

Jensen-Shannon divergence similarity (JSDs) was used to quantify network edges which represented the interregional similarity in the distributions of cortical thickness.^25^ ^26^ Specifically, values of all vertices within each region were first extracted, based on which a probability density function was estimated using a kernel density function (MATLAB function, ksdensity). After converting the probability density functions into probability distributions (PDs), the JSDs between each pair of ROIs was calculated as^26^:

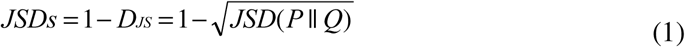

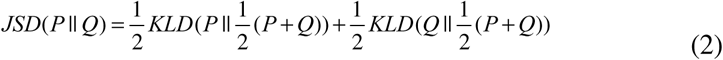

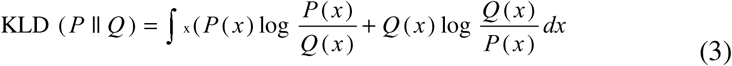

where P and Q presented regional PDs. The value range for JSDs was [0, 1], with 0 denoting absolutely different and 1 exactly the same PDs.

### Site effects correction

Multiple sites were included in this study. We applied a ComBat harmonization to correct the site effects on morphological similarities before the following statistical analysis.^27^ ^28^ ComBat harmonization was applied to remove site-related mean and variance effects while preserving biological variability.^29^

### Graph theoretical analysis

For the constructed single-subject MBNs, their topological properties were quantified with the GRETNA toolbox.^30^

### Graph theory-based network analysis

To identify alterations in network topological characteristics in ASD patients, we calculated five nodal centrality, including degree centrality, PageRank centrality, betweenness centrality, eigenvector centrality and efficiency. In addition, we also calculated four global metrics, including characteristic path length (*lp*), normalized *lp,* clustering coefficient (*cp*), and normalized *cp*. Detailed formulas and interpretations of these measures can be found elsewhere.^31^

### Selection of threshold

Before topologically characterizing the MBNs derived above, a consecutive sparsity ranging from 0.026 to 0.4 with an interval of 0.02 was used to convert the MBNs to binary sparse networks. To achieve networks with estimable small-world attributes, the sparsity range was meticulously selected to ensure that the networks remained sparse, as established in previous studies.^32^ ^33^ Additionally, to prevent the formation of isolated nodes or the presence of multiple disconnected components within these networks, we incorporated a minimum spanning tree algorithm^34^into the sparsity-based thresholding process. This integration was crucial for maintaining the structural integrity and connectivity of the resulting networks.

### Area under curve (AUC) value

AUC values under the range of sparsity, which provided a summary scalar for the topological characterization of brain networks, were calculated for the network measures for statistical analysis. The AUC was computed for each network measure Y and expressed as follows:

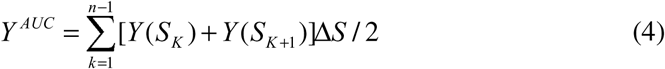

where sparsity thresholds {S_k_| k = 1,2,…,n} ranged from S_1_ = 0.026 to S_n_ = 0.4 with an interval ΔS = 0.02.

## Statistical analysis

### Demographic and clinical variables

The statistical analysis was carried out using SPSS software (version 25.0) concerning the demographic and clinical traits of the two groups. Two-sample t-test was used to examine the differences of continuous variables in demographic and neuropsychological data between ASD and TD groups. Chi-square test was used for the categorical variables.

### Network measures

For all global and nodal network metrics, between-group differences were statistically tested with nonparametric permutation tests (10,000 times) controlling for age and IQ as covariates. To address multiple comparisons, false discovery rate (FDR) correction was applied across all regions for each nodal measure (q<0.05). Effect sizes (Cohen’s d) were calculated for all significant findings.

### Morphological similarity

Based on the findings that nodal metrics of the lateral area of left OFC altered in the ASD group (see results), we further examined the differences of morphological similarity between the left lateral OFC and the other regions using nonparametric permutation tests based on the t statistics derived from two-sample t-tests (10,000 times). Specifically, age and IQ were treated as covariates in order to exclude their potential effects. The FDR procedure was used to correct for multiple comparisons across 207 regions at the level of *q* < 0.05. Furthermore, we tested whether the altered morphological similarity linked to the lateral area of left OFC was susceptible to particular structural lobes (frontal, temporal, parietal, insular, limbic, and occipital lobes) or functional subnetworks (visual, somatomotor, dorsal attention, ventral attention, limbic, frontoparietal and default mode subnetworks). Briefly, we firstly recorded the frequencies of the brain regions showing altered morphological similarity linked to the lateral area of left OFC in ASD in each structural lobe and functional subnetwork. Then, we randomly selected the same number of brain regions and recorded their frequencies in each structural lobe and functional subnetwork. After 10,000 repetitions, a null distribution was obtained for each structural lobe and functional subnetwork, based on which a *p-*value was calculated as the proportion of random times that yielded equal or greater frequencies than the actual observation. The FDR procedure was used to correct for multiple comparisons across six structural lobes or seven functional subnetworks at the level of *q* < 0.05.

### Association between morphological similarity and regional neurotransmitter systems

To examine the molecular correlates of cortical morphological networks, we utilized the JuSpace database^35^ (https://github.com/juryxy/JuSpace), which provides PET-derived neurotransmitter receptor and transporter distribution maps obtained from healthy populations. A total of 27 maps were initially considered, including those associated with cannabinoid (CB1), dopaminergic (D1, D2, DAT, 18F-DOPA), GABAergic (GABAa), glutamatergic (mGluR5), opioid (μ-opioid), noradrenergic (NAT), serotonergic (SERT, 5-HT1a, 5-HT1b, 5-HT2a, 5-HT4), and cholinergic (VAChT) systems. Due to known issues with cortical binding fidelity, the D2 receptor map (using the raclopride tracer) was excluded, yielding a final set of 26 neurochemical maps for analysis. All neurotransmitter maps were aligned to the same brain atlas used in morphological similarity analysis, ensuring spatial correspondence. The molecular analytic pipeline involved three primary steps: (1) extraction of regional mean signal intensities for each neurotransmitter map across all predefined ROIs; and (2) Z-score normalization of each intensity profile to enhance comparability across different neurotransmitter systems.

We implemented partial least-squares (PLS) regression to evaluate the relationship between each neurotransmitter’s regional intensity pattern and the OFC-based connectivity vector. Spatial autocorrelation was accounted for by performing 10,000 iterations of spin rotation randomization, generating empirical null distributions for significance estimation. Correction for multiple comparisons across neurotransmitter measures was conducted using the false discovery rate (FDR) procedure with a threshold of q<0.05. To further assess whether specific neurotransmitter systems contributed to the observed correlation, we extracted the weight of each neurotransmitter map to the first component of PLS (PLS1) score and calculated the Z-score by bootstrapping test (10,000 times). The neurotransmitter maps with absolute Z-score higher than 1.96 were considered contributing to the observed correlation significantly.

### Clinical relationship between Autism Diagnostic Observation Schedule (ADOS) and morphological similarities

In addition, an exploratory analysis was conducted to examine the association between OFC-related morphological similarity and ADOS total scores in ASD participants.

### Reproducibility analysis

A series of analysis was conducted to evaluate the reproducibility of graph metrics. First, To examine the robustness of findings, we constructed weighted MBNs using the same similarity matrices. Second, bootstrap analysis, a widely used statistical method for assessing the robustness, bias, and variability of numerical estimates, was employed to mitigate the influence of individual-level variability.^36^ In each bootstrap iteration, we randomly selected 90% of participants and computed the mean value of each metric within this subsample. This process was repeated 10,000 times. The coefficient of variation (CV) across these iterations was calculated to quantify metric stability.

## Results

### The demographic and psychiatric characteristics of participants

The demographic and psychiatric measurements are presented in **Table 1**. Age and handedness showed no significant differences between the two groups (age: t=-0.21, *p*=0.840; handedness: t=1.02, *p*=0.800). As for IQ, the score of full scale IQ and subscore of verbal IQ were significantly lower in the ASD group than those in TD group (full scale IQ: t=-3.08, *p*=0.002; verbal IQ: t=-4.79, *p*<0.001). There was no significant difference between the two groups in the score of performance IQ.

### Global and nodal topological metrics of MBNs between adolescent males with ASD and TD

For global metrics, adolescent males with ASD exhibited significantly increased normalized *cp* compared with TD (t=2.40, *p*=0.020, FDR corrected, Cohen’s *d*=0.23) (**Figure 2A**). No significant differences between the two groups were observed in *cp* (t=0.81, *p*=0.41, Cohen’s d=0.10), *lp* (t=-0.87, *p*=0.39), or normalized *lp* (t=-0.16, *p*=0.87).

**Figure 2.**
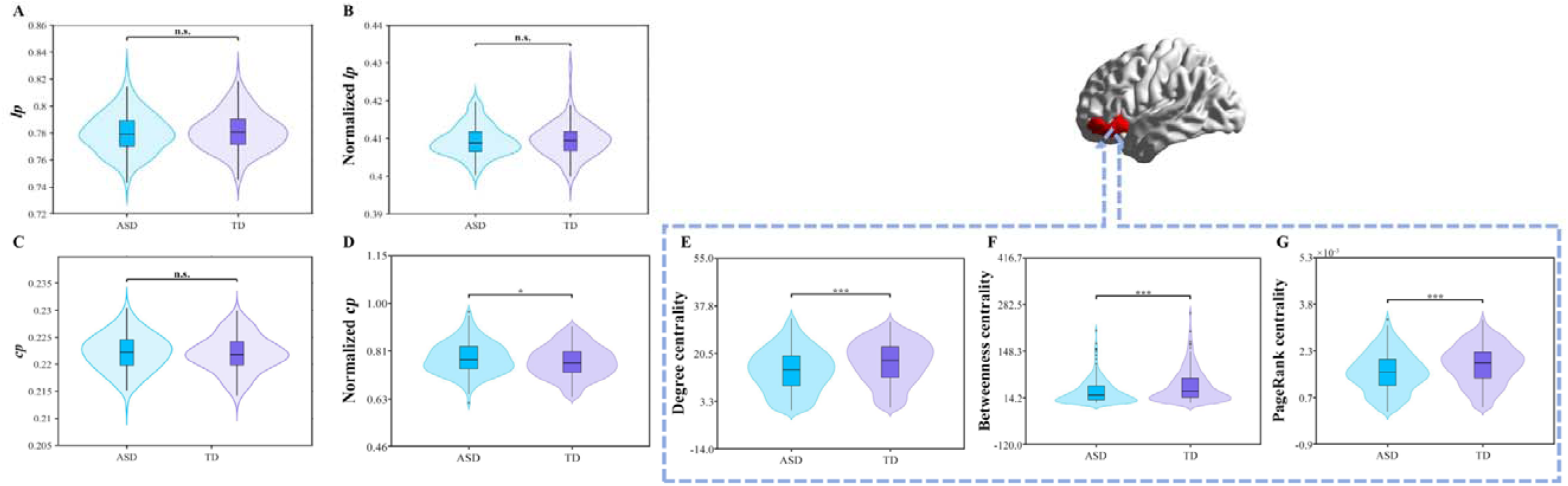
The differences of global metrics and nodal centrality of left lateral OFC between adolescent males with ASD and TD. (A) *lp*; (B) Normalized *lp*; (C) *cp*; (D) Normalized *cp*; (E) Degree centrality of the left lateral OFC; (F) Betweenness centrality of the left lateral OFC; (G) PageRank centrality of the left lateral OFC; ASD, autism spectrum disorder; TD, typically developing; OFC, orbitofrontal cortex; lp, characteristic path length; cp, clustering coefficient; ***, *p*<0.001; *, *p*<0.05; n.s., not significant.

For nodal metrics, adolescent males with ASD showed altered nodal topological metrics only in left lateral OFC. Specifically, adolescent males with ASD showed significant decreases in nodal degree centrality (t=-3.99, *p*<0.001, FDR corrected, Cohen’s *d*=0.39), PageRank centrality (t=-3.92, *p*<0.001, FDR corrected, Cohen’s *d*=0.39) and betweenness centrality (t=-4.04, *p*<0.001, FDR corrected, Cohen’s *d*=0.40) (**Figure 2B**). No significant differences were observed in either eigenvector centrality or efficiency between the two groups.

### Differences in morphological similarity

Given that the altered nodal metrics among adolescent males with ASD were observed in the lateral area of left OFC, we further examined the differences of morphological similarity linked to the lateral area of left OFC between ASD and TD. We identified decreased morphological similarities between lateral area of left OFC and 65 ROIs in ASD compared with TD (**Figure 3**). With respect to the 65 ROIs, we analyzed them from the perspective of structural lobes and functional subnetworks, respectively. We found that the ROIs showing altered morphological similarity linked to the lateral area of left OFC were inclined to be located in the frontal lobe (27/68, *p*=0.049) and dorsal attention subnetwork (14/30, *p*=0.043).

**Figure 3.**
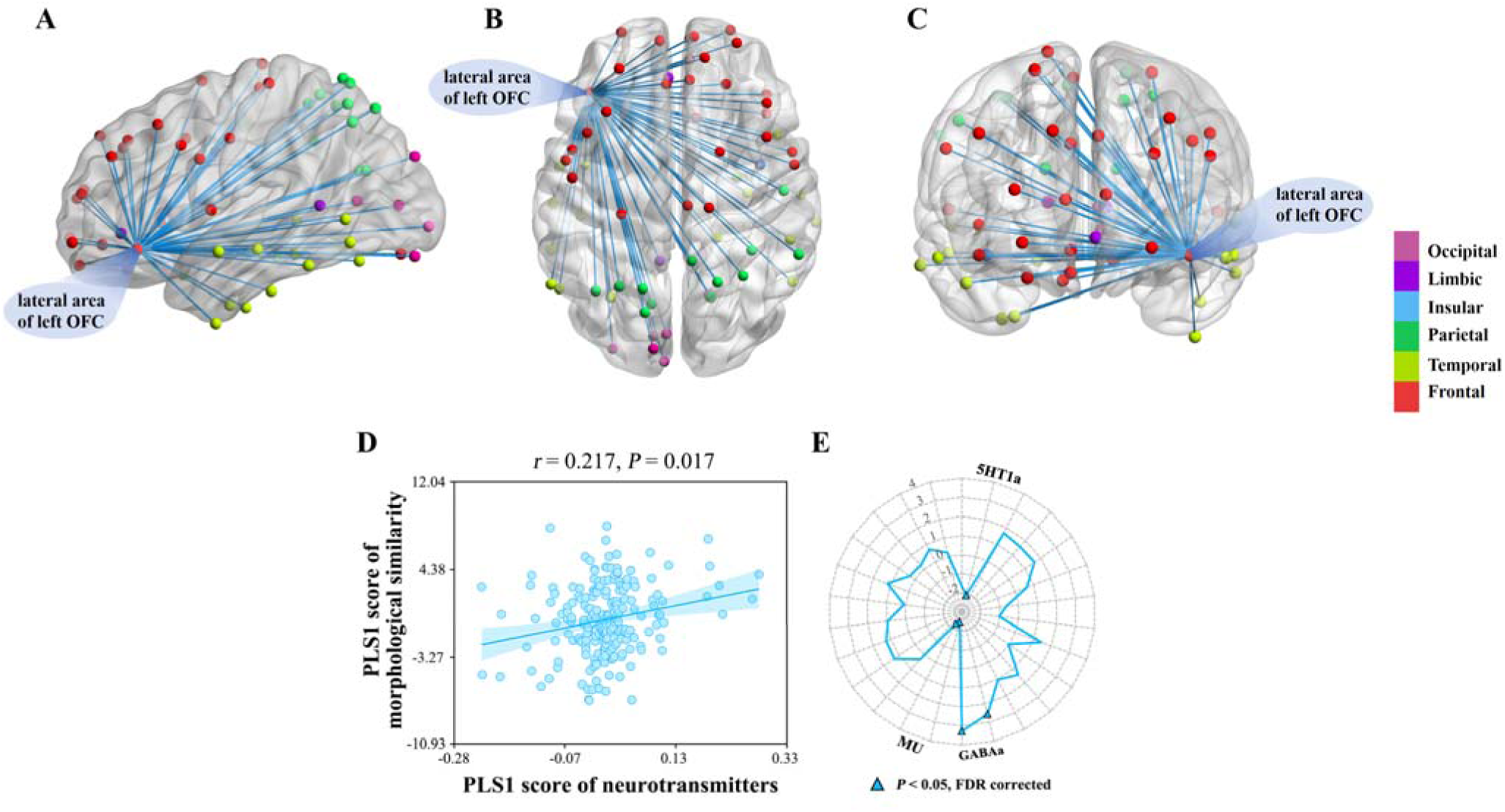
Reduced morphological similarity of the left lateral OFC in adolescent males with ASD and its associations with neurotransmitter distributions. (A-C) Visualization of reduced morphological similarity between the left lateral OFC and other cortical regions in adolescent males with ASD, displayed from (A) lateral view; (B) dorsal view; (C) medial views. (D) correlation analysis showing a significant positive association between morphological similarity and neurotransmitter densities (r=0.217, p=0.017). (E) Neurotransmitter contribution weights to PLS1, with significant effects observed for 5-HT1a, GABAa, and μ-opioid receptor systems. (p<0.05, FDR corrected). ASD, autism spectrum disorder; TD, typically developing; OFC, orbitofrontal cortex; PLS1, the first component of the partial least-squares model; FDR, false discovery rate.

### Multivariate modeling revealed significant molecular association with left lateral OFC morphological similarity

We applied PLSR model to assess how well the spatial distributions of 26 neurotransmitter receptor and transporter maps jointly explain the whole-brain morphological similarity profile of the left lateral OFC. The first latent component explained 22.7% of the total variance (R_²_=0.227) and showed a significant multivariate correspondence (r =0.48, p=0.017, spin-corrected). After transforming the weights of PLS1 score to Z-score, five neurotransmitter maps were considered to contribute significantly to the PLS1, including 5-HT1a (Z=-2.09, *p*=0.036), GABAa (Z=2.55, *p*=0.011,^35^; Z=3.30, *p*<0.001,^37^), and μ-opioid (Z=-2.45, *p*=0.014,^38^; Z=-2.32, *p*=0.021,^39^) (**Figure 4**).

**Figure 4.**
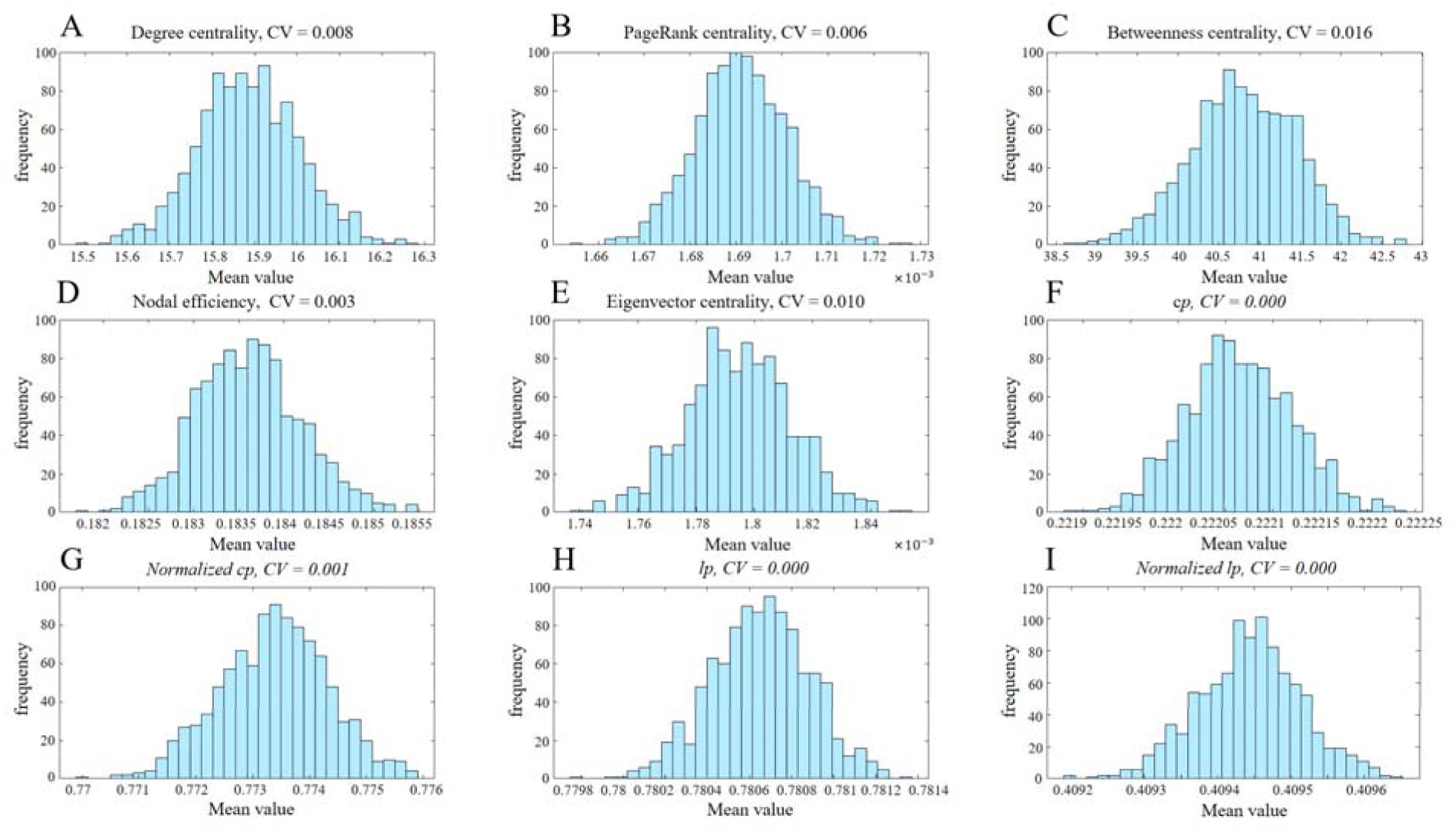
Bootstrap distributions of graph-theoretical metrics derived from MBNs. Histograms show the resampled mean values and corresponding CV for (A) Degree centrality; (B) PageRank centrality; (C) Betweenness centrality; (D) Nodal efficiency; (E) Eigenvector centrality; (F) *cp*; (G) Normalized *cp*; (H) *lp*; and (I) Normalized *lp*. The narrow distributions and low CV values indicate the robustness and stability of these graph metrics under bootstrap resampling. CV: coefficient of variation; *cp*: clustering coefficient; *lp*: characteristic path length.

### Clinical relationship between ADOS and morphological similarity

An exploratory analysis was conducted to examine the association between OFC-related morphological similarity and ADOS total scores in ASD participants. However, no significant association was observed (R_²_=0.023, *p*=0.752).

### Reproducibility analysis results

To examine the robustness of findings, we constructed weighted MBNs using the same matrices. Crucially, the key alteration in the left lateral OFC was robustly replicated under a weighted network framework. While the specific suite of significant nodal metrics differed slightly—with weighted networks showing significance in PageRank centrality, degree centrality, and efficiency, and binarized networks in degree centrality, PageRank centrality, and betweenness centrality—the convergence on the left lateral OFC across both methodologies was striking. To further interrogate this consistent node-level effect, we examined its morphological connectivity pattern. We found a substantial and highly comparable reduction in the number of connecting edges to surrounding regions in both the binarized (65 edges reduced) and weighted (62 edges reduced) networks. Together, these multi-faceted validation analyses strongly underscore the reliability and reproducibility of the functional disruption identified in the left lateral OFC. These results were reported in the supplementary materials (**Figure S2, S3**).

In addition, bootstrap analysis demonstrated very low CV across all metrics, underscoring robustness to individual-level variability. For global metrics, CV were <0.001 for *cp*, normalized *cp*, *lp*, and normalized *lp*. For nodal metrics within the left lateral OFC, CV ranged from 0.003 to 0.016 across degree centrality, PageRank centrality, betweenness centrality, efficiency centrality, and eigenvector centrality (**Figure 5**), further confirming the stability of these estimates.

## Discussions

### Main findings

This study reveals significant disruptions in single-subject MBNs among adolescent males with ASD, with the left lateral OFC emerging as a key hub of dysorganization. We identified three core abnormalities: (i) In terms of topological metrics, adolescent males with ASD exhibited a distinct network profile characterized by an increased normalized *cp* and reduced nodal measures in the left lateral OFC specifically degree, PageRank, and betweenness centrality; (ii) The left lateral OFC had reduced morphological similarity, mainly with frontal and dorsal attention regions; (iii) OFC morphological similarity were significantly associated with serotonin (5-HT1a), GABAa, and μ-opioid receptor distributions. Together, these findings identify the lateral OFC as a key network hub linking structural alterations to neurochemical systems, suggesting a potential target for future neuromodulation strategies.

The rationale for employing graph-based centrality metrics stems from their ability to quantify individual nodes’ importance and influence within a network^40^ thereby reflecting their hub role in information transfer and integration.^31^ In this study, we observed that adolescent males with ASD exhibited significant reductions across multiple centrality indices of the left lateral OFC, including nodal degree, PageRank, and betweenness centrality. This consistent finding indicates that the hub status of the OFC was weakened, with a marked decline in its role in cross-regional information transfer and global integration. In line with previous research, the OFC has been regarded as a critical hub for emotional regulation, social reward processing, and decision-making, and its dysfunction has been shown to be closely associated with social communication deficits and behavioral inflexibility in ASD.^41–43^ Therefore, the nodal-level results provide direct structural network evidence for the symptomatic manifestations of ASD, suggesting that OFC hub dysfunction may constitute a key pathological mechanism of the disorder.

More importantly, these nodal abnormalities were not isolated but were accompanied by significant reductions in morphological similarities between the left lateral OFC and 65 other regions. In other words, left lateral OFC hub anomalies co-occurred with widespread edge-level disruptions, revealing a multi-level abnormal pattern of “nodal dysfunction plus weakened morphological connectivity.” This dual abnormality further reinforces the dysregulated hub status of the OFC within the ASD structural connectome. Previous large-sample structural studies have similarly reported reductions in OFC gray matter volume and aberrant structural covariance patterns in individuals with ASD.^44–46^ Our findings replicate and extend these observations within an individualized MBN framework, providing more integrative evidence for the structural network pathology of ASD.

This convergent evidence is not only of anatomical significance but also suggests potential functional implications. The OFC plays a central role in social-emotional processing,^41^ reward valuation,^42^ and behavioral flexibility,^43^ domains in which individuals with ASD consistently exhibit impairments. Therefore, OFC hub abnormalities, together with widespread reductions in cross-regional morphological connections, may jointly contribute to deficits in social motivation, emotional regulation, and behavioral flexibility in ASD. In other words, the OFC, as a “dysfunctional epicenter” within the ASD structural connectome, represents not only an abnormal node at the topological level but also a critical source of global network imbalance. Notably, this multi-level abnormal pattern was consistently replicated within the weighted MBN framework. Bootstrap robustness analyses further corroborated these results, supporting their reliability and stability.

Despite being derived from healthy individuals, the neurotransmitter atlases have demonstrated validity across disorders. For example, Dukart et al.^35^ found that Parkinson’s disease MRI changes coincide with normative topographies of dopamine D2. Similarly, Premi et al.^47^ used JuSpace to identify distinct neurochemical patterns in genetic frontotemporal dementia (FTD), and extended the approach to primary progressive aphasia, again linking transmitter maps to gray-matter alterations.^48^ Tang et al. (2024)^49^ further showed significant correlations between normative receptor distributions and structural atrophy in Alzheimer’s disease, while Kang et al. (2024)^50^ corroborated these findings through multimodal meta-analysis. Together, these studies underscore the methodological robustness and practical value of JuSpace’s normative PET-based neurotransmitter maps for investigating structural–neurochemical relationships in disease contexts.

The μ-opioid system has been implicated in behavioral regulation, particularly in relation to impulsivity.^51^ Large-scale population-based research (>44,000 children) has shown that autism-related traits often co-occur with hyperactive-impulsive behaviors, highlighting impulsivity as a potential behavioral dimension of ASD and supporting investigation of related neurobiological systems such as the μ-opioid pathway.^52^ In the present study, we found that this morphological connectivity pattern was associated with the intrinsic distribution of the μ-opioid pathway in adolescent males with ASD. The prefrontal cortex (PFC) contains appreciable densities of μ-opioid receptors, and μ-opioid receptor signaling can modulate PFC neuronal activity.^53^ Excessive opioid signaling may impair inhibitory control by altering the executive and motivational functions of the PFC.^53^ In animal models, genetically modified mice lacking μ-opioid receptors exhibit markedly reduced motor impulsivity, suggesting a role for μ-opioid receptors in promoting impulsive behavior.^54^ Together, these findings indicate that μ-opioid signaling could be involved in the molecular mechanisms underlying ASD and provide supporting evidence for considering opioid antagonists as potential pharmacological interventions.

In the present study, we found that this morphological similarity was positively correlated with GABAa receptor distribution. Consistent with this, PET studies have reported reduced α5 GABAa receptor levels in individuals with ASD,^55^ suggesting that deficits in GABAa receptor availability could influence OFC network integration. Supporting evidence from postmortem studies has shown reduced expression of GABAa receptor subunits α4, α5, and β1, as well as the GABAb receptor subunit β1 in ASD,^56^ reinforcing the hypothesis that GABA system dysregulation plays an important role in ASD pathophysiology. Given the recognized significance of E/I imbalance in ASD, elucidating the specific functions of individual GABAa receptor subtypes in animal models could facilitate the development of subtype-selective modulators as promising therapeutic strategies.^57^ The present study provides structural network evidence to inform future research in this direction.

In addition, we also found that this morphological similarity was positively correlated with the distribution of 5-HT1a receptors. Similarly, our group previously has reported that white matter morphological networks is significantly associated with the spatial distribution of 5-HT1a receptors.^58^ Evidence from animal studies has shown that activation of 5-HT1a receptors can modulate axonal excitability in the dorsal columns of the rat spinal cord,^59^ indicating an important role of 5-HT1a signaling in regulating neural conduction properties. In human studies, infants exposed to selective serotonin reuptake inhibitors have been found to exhibit increased white matter structural connectivity.^60^ Taken together, these cross-species and cross-modality findings support the notion that the intrinsic distribution pattern of 5-HT1a receptors may not only influence the structural organization of white matter networks but also shape the morphological similarity features of gray matter. Our findings provide correlation-based evidence from the morphological gray matter network perspective for the potential role of 5-HT1a in network integration.

### Limitations

Our use of single-subject MBNs via Jensen-Shannon divergence advances beyond traditional group-level structural covariance analysis,^25^ enabling detection of individual-specific network pathologies for personalized biomarkers.^7^ ^61^ Despite the novel insights provided by this study, several limitations should be acknowledged. Firstly, the current analysis focused exclusively on male adolescents with ASD, owing to the demographic composition of the ABIDE cohort. As ASD presents with sex-specific neurodevelopmental trajectories and phenotypic variability,^62^ future studies are needed to determine whether similar morphological network alterations are present in females. Secondly, although psychotropic medications are known to influence cortical morphology and network properties, detailed medication histories were not consistently available in the current dataset, preventing a systematic assessment of their potential confounding effects. Thirdly, the morphological brain networks (MBNs) were constructed solely based on cortical thickness similarity. While this approach captures coordinated maturational or pathological processes, it does not directly reflect white matter connectivity. Integrating multi-modal data, such as diffusion imaging or functional MRI, could offer a more comprehensive characterization of ASD-related network alterations. Fourth, normative PET maps used in this study were derived from healthy adults. While these templates may not fully capture adolescent-specific neurotransmitter distributions, previous validations suggest their utility in disease-related structural studies. Nonetheless, future work incorporating adolescent- or ASD-specific PET data will be essential to further refine the neurochemical interpretation of our findings.

### Implications

Adolescence is a critical and vulnerable stage for prefrontal maturation, marked by regionally asynchronous processes such as synaptic pruning, dendritic arborization, and intracortical myelination that reshape brain connectivity.^63^ ^64^ The OFC, with its protracted developmental trajectory, is particularly sensitive to such reorganization. Our findings indicate that adolescent males with ASD exhibit convergent OFC-centered morphological similarity, including reduced hub centrality (degree centrality, PageRank centrality, betweenness centrality) and weakened morphological similarity across 65 edges. These abnormalities may reflect disrupted normative refinement (e.g., aberrant pruning or myelination within OFC circuits)^65^ and an increased susceptibility to “second hits” during adolescence, when hormonal and environmental influences interact with extended OFC plasticity.^66^ Notably, OFC dysfunction intensifies during adolescence when advanced social-emotional regulation is required.^67^ The male predominance observed here highlights sex-specific trajectories, as testosterone and estrogen differentially affect prefrontal myelination and GABAergic signaling, potentially exacerbating OFC disconnectivity in males.^68–70^ Collectively, these findings suggest that adolescence constitutes a significant window in which disrupted maturational mechanisms consolidate into persistent network dysfunction, underscoring puberty as a pivotal period for targeted interventions such as GABAergic modulation or cognitive remediation.

## Conclusion

These findings highlight the left lateral OFC as a structural key hub in adolescent males with ASD, linking left lateral OFC-based morphological similarity to neurotransmitter systems and providing a potential neurobiological basis for targeted interventions in this population.

## Data Availability

Ethics approval was acquired for the original studies in the ABIDE repository.

http://fcon_1000.projects.nitrc.org/indi/abide/

## Acknowledgements

We would like to thank the contributors and organizers of the ABIDE-I and II datasets.

## Contributors

X.W. and J.W. provided guidance and advice; X.W. and J.W. designed research; H.Z., J.L. and C.H. collected the data; H.Z.,C.H. , Y.H., L.M. and B.X. performed research; H.Z. and J.L. drafted the manuscript.

## Patient consent for publication

Not applicable.

## Ethics approval

Ethics approval was acquired for the original studies in the ABIDE repository.

## Provenance and peer review

Not commissioned; externally peer reviewed.

## Data availability statement

The datasets analyzed during the current study are available from the corresponding author on reasonable request.

## REFERENCES

1 American Psychiatric Association. Diagnostic and statistical manual of mental disorders: DSM-5. American Psychiatric Association, 2013.

2 Jeste SS and Geschwind DH. Disentangling the heterogeneity of autism spectrum disorder through genetic findings. Nat Rev Neurol 2014;10:74–81.

3 Bedford SA, Park MTM, Devenyi GA, et al. Large-scale analyses of the relationship between sex, age and intelligence quotient heterogeneity and cortical morphometry in autism spectrum disorder. Mol Psychiatry 2020;25:614–28.

4 Raj A, Mueller SG, Young K, et al. Network-level analysis of cortical thickness of the epileptic brain. Neuroimage 2010;52:1302–13.

5 Tijms BM, Seriès P, Willshaw DJ, et al. Similarity-based extraction of individual networks from gray matter MRI scans. Cereb Cortex 2012;22:1530–41.

6 Zhou L, Wang Y, Li Y, et al. Hierarchical anatomical brain networks for MCI prediction: revisiting volumetric measures. PLoS One 2011;6:e21935.

7 He C, Cortes JM, Kang X, et al. Individual-based morphological brain network organization and its association with autistic symptoms in young children with autism spectrum disorder. Hum Brain Mapp 2021;42:3282–94.

8 Gao J, Chen M, Li Y, et al. Multisite Autism Spectrum Disorder Classification Using Convolutional Neural Network Classifier and Individual Morphological Brain Networks. Front Neurosci 2020;14:629630.

9 Bilgen I, Guvercin G and Rekik I. Machine learning methods for brain network classification: Application to autism diagnosis using cortical morphological networks. J Neurosci Methods 2020;343:108799.

10 Soussia M and Rekik I. Unsupervised Manifold Learning Using High-Order Morphological Brain Networks Derived From T1-w MRI for Autism Diagnosis. Front Neuroinform 2018;12:70.

11 Zielinski BA, Prigge MB, Nielsen JA, et al. Longitudinal changes in cortical thickness in autism and typical development. Brain 2014;137:1799–812.

12 Romero-Garcia R, Warrier V, Bullmore ET, et al. Synaptic and transcriptionally downregulated genes are associated with cortical thickness differences in autism. Mol Psychiatry 2019;24:1053–64.

13 Wallace GL, Eisenberg IW, Robustelli B, et al. Longitudinal cortical development during adolescence and young adulthood in autism spectrum disorder: increased cortical thinning but comparable surface area changes. J Am Acad Child Adolesc Psychiatry 2015;54:464–9.

14 Fiore A, Preziosa P, Tedone N, et al. Correspondence among gray matter atrophy and atlas-based neurotransmitter maps is clinically relevant in multiple sclerosis. Mol Psychiatry 2023;28:1770–82.

15 Ren J, Yan L, Zhou H, et al. Unraveling neurotransmitter changes in de novo GBA-related and idiopathic Parkinson’s disease. Neurobiol Dis 2023;185:106254.

16 Morais RF, Sousa JM, Koba C, et al. Differential involvement of neurotransmitter pathways in AD, bvFTD and MCI: Whole-brain MRI analysis. Neurobiol Dis 2025;209:106897.

17 Di Martino A, Yan CG, Li Q, et al. The autism brain imaging data exchange: towards a large-scale evaluation of the intrinsic brain architecture in autism. Mol Psychiatry 2014;19:659–67.

18 Uddin LQ, Supekar K, Lynch CJ, et al. Salience network-based classification and prediction of symptom severity in children with autism. JAMA Psychiatry 2013;70:869–79.

19 Tohka J, Zijdenbos A and Evans A. Fast and robust parameter estimation for statistical partial volume models in brain MRI. Neuroimage 2004;23:84–97.

20 Dahnke R, Yotter RA and Gaser C. Cortical thickness and central surface estimation. Neuroimage 2013;65:336–48.

21 Yotter RA, Thompson PM and Gaser C. Algorithms to improve the reparameterization of spherical mappings of brain surface meshes. J Neuroimaging 2011;21:e134–47.

22 Yotter RA, Dahnke R, Thompson PM, et al. Topological correction of brain surface meshes using spherical harmonics. Hum Brain Mapp 2011;32:1109–24.

23 Ashburner J. A fast diffeomorphic image registration algorithm. Neuroimage 2007;38:95–113.

24 Fan L, Li H, Zhuo J, et al. The Human Brainnetome Atlas: A New Brain Atlas Based on Connectional Architecture. Cereb Cortex 2016;26:3508–26.

25 Alexander-Bloch A, Giedd JN and Bullmore E. Imaging structural co-variance between human brain regions. Nat Rev Neurosci 2013;14:322–36.

26 Li Y, Wang N, Wang H, et al. Surface-based single-subject morphological brain networks: Effects of morphological index, brain parcellation and similarity measure, sample size-varying stability and test-retest reliability. Neuroimage 2021;235:118018.

27 Achard S, Salvador R, Whitcher B, et al. A resilient, low-frequency, small-world human brain functional network with highly connected association cortical hubs. J Neurosci 2006;26:63–72.

28 Johnson WE, Li C and Rabinovic A. Adjusting batch effects in microarray expression data using empirical Bayes methods. Biostatistics 2007;8:118–27.

29 Fortin JP, Cullen N, Sheline YI, et al. Harmonization of cortical thickness measurements across scanners and sites. Neuroimage 2018;167:104–20.

30 Wang J, Wang X, Xia M, et al. GRETNA: a graph theoretical network analysis toolbox for imaging connectomics. Front Hum Neurosci 2015;9:386.

31 Rubinov M and Sporns O. Complex network measures of brain connectivity: uses and interpretations. Neuroimage 2010;52:1059–69.

32 Wang J, Wang L, Zang Y, et al. Parcellation-dependent small-world brain functional networks: a resting-state fMRI study. Hum Brain Mapp 2009;30:1511–23.

33 Watts DJ and Strogatz SH. Collective dynamics of ’small-world’ networks. Nature 1998;393:440–2.

34 Tewarie P, van Dellen E, Hillebrand A, et al. The minimum spanning tree: an unbiased method for brain network analysis. Neuroimage 2015;104:177–88.

35 Dukart J, Holiga S, Rullmann M, et al. JuSpace: A tool for spatial correlation analyses of magnetic resonance imaging data with nuclear imaging derived neurotransmitter maps. Hum Brain Mapp 2021;42:555–66.

36 Lemoine F, Domelevo Entfellner JB, Wilkinson E, et al. Renewing Felsenstein’s phylogenetic bootstrap in the era of big data. Nature 2018;556:452–56.

37 Nørgaard M, Beliveau V, Ganz M, et al. A high-resolution in vivo atlas of the human brain’s benzodiazepine binding site of GABA(A) receptors. Neuroimage 2021;232:117878.

38 Kantonen T, Karjalainen T, Isojärvi J, et al. Interindividual variability and lateralization of μ-opioid receptors in the human brain. Neuroimage 2020;217:116922.

39 Nummenmaa L, Jern P, Malén T, et al. μ-opioid receptor availability is associated with sex drive in human males. Cogn Affect Behav Neurosci 2022;22:281–90.

40 Ille S, Zhang H, Stassen N, et al. Noninvasive- and invasive mapping reveals similar language network centralities - A function-based connectome analysis. Cortex 2024;174:189–200.

41 Zalla T and Sperduti M. The amygdala and the relevance detection theory of autism: an evolutionary perspective. Front Hum Neurosci 2013;7:894.

42 O’Doherty J, Kringelbach ML, Rolls ET, et al. Abstract reward and punishment representations in the human orbitofrontal cortex. Nat Neurosci 2001;4:95–102.

43 D’Cruz AM, Ragozzino ME, Mosconi MW, et al. Reduced behavioral flexibility in autism spectrum disorders. Neuropsychology 2013;27:152–60.

44 Sha Z, van Rooij D, Anagnostou E, et al. Subtly altered topological asymmetry of brain structural covariance networks in autism spectrum disorder across 43 datasets from the ENIGMA consortium. Mol Psychiatry 2022;27:2114–25.

45 Mei T, Llera A, Floris DL, et al. Gray matter covariations and core symptoms of autism: the EU-AIMS Longitudinal European Autism Project. Mol Autism 2020;11:86.

46 Ni HC, Lin HY, Chen YC, et al. Boys with autism spectrum disorder have distinct cortical folding patterns underpinning impaired self-regulation: a surface-based morphometry study. Brain Imaging Behav 2020;14:2464–76.

47 Premi E, Pengo M, Mattioli I, et al. Early neurotransmitters changes in prodromal frontotemporal dementia: A GENFI study. Neurobiol Dis 2023;179:106068.

48 Premi E, Dukart J, Mattioli I, et al. Unravelling neurotransmitters impairment in primary progressive aphasias. Hum Brain Mapp 2023;44:2245–53.

49 Tang X, Guo Z, Chen G, et al. A Multimodal Meta-Analytical Evidence of Functional and Structural Brain Abnormalities Across Alzheimer’s Disease Spectrum. Ageing Res Rev 2024;95:102240.

50 Kang X, Wang D, Lin J, et al. Convergent Neuroimaging and Molecular Signatures in Mild Cognitive Impairment and Alzheimer’s Disease: A Data-Driven Meta-Analysis with N = 3,118. Neurosci Bull 2024;40:1274–86.

51 Giacomini JL, Geiduschek E, Selleck RA, et al. Dissociable control of μ-opioid-driven hyperphagia vs. food impulsivity across subregions of medial prefrontal, orbitofrontal, and insular cortex. Neuropsychopharmacology 2021;46:1981–89.

52 D’Urso S, Moen GH, Hwang LD, et al. Intrauterine Growth and Offspring Neurodevelopmental Traits: A Mendelian Randomization Analysis of the Norwegian Mother, Father and Child Cohort Study (MoBa). JAMA Psychiatry 2024;81:144–56.

53 Baldo BA. Prefrontal Cortical Opioids and Dysregulated Motivation: A Network Hypothesis. Trends Neurosci 2016;39:366–77.

54 Olmstead MC, Ouagazzal AM and Kieffer BL. Mu and delta opioid receptors oppositely regulate motor impulsivity in the signaled nose poke task. PLoS One 2009;4:e4410.

55 Mendez MA, Horder J, Myers J, et al. The brain GABA-benzodiazepine receptor alpha-5 subtype in autism spectrum disorder: a pilot [(11)C]Ro15-4513 positron emission tomography study. Neuropharmacology 2013;68:195–201.

56 Fatemi SH, Reutiman TJ, Folsom TD, et al. mRNA and protein levels for GABAAalpha4, alpha5, beta1 and GABABR1 receptors are altered in brains from subjects with autism. J Autism Dev Disord 2010;40:743–50.

57 Rudolph U and Möhler H. GABAA receptor subtypes: Therapeutic potential in Down syndrome, affective disorders, schizophrenia, and autism. Annu Rev Pharmacol Toxicol 2014;54:483–507.

58 Li J, Jin S, Li Z, et al. Morphological Brain Networks of White Matter: Mapping, Evaluation, Characterization, and Application. Adv Sci (Weinh) 2024;11:e2400061.

59 Saruhashi Y, Young W and Hassan AZ. Calcium-mediated intracellular messengers modulate the serotonergic effects on axonal excitability. Neuroscience 1997;81:959–65.

60 Lugo-Candelas C, Cha J, Hong S, et al. Associations Between Brain Structure and Connectivity in Infants and Exposure to Selective Serotonin Reuptake Inhibitors During Pregnancy. JAMA Pediatr 2018;172:525–33.

61 Wang H, Jin X, Zhang Y, et al. Single-subject morphological brain networks: connectivity mapping, topological characterization and test-retest reliability. Brain Behav 2016;6:e00448.

62 Lai MC, Lombardo MV and Baron-Cohen S. Autism. Lancet 2014;383:896–910.

63 Gogtay N, Giedd JN, Lusk L, et al. Dynamic mapping of human cortical development during childhood through early adulthood. Proc Natl Acad Sci U S A 2004;101:8174–9.

64 Mills KL, Goddings AL, Herting MM, et al. Structural brain development between childhood and adulthood: Convergence across four longitudinal samples. Neuroimage 2016;141:273–81.

65 Johnson CM, Loucks FA, Peckler H, et al. Long-range orbitofrontal and amygdala axons show divergent patterns of maturation in the frontal cortex across adolescence. Dev Cogn Neurosci 2016;18:113–20.

66 Frere PB, Vetter NC, Artiges E, et al. Sex effects on structural maturation of the limbic system and outcomes on emotional regulation during adolescence. Neuroimage 2020;210:116441.

67 Supekar K, Uddin LQ, Khouzam A, et al. Brain hyperconnectivity in children with autism and its links to social deficits. Cell Rep 2013;5:738–47.

68 Crone EA and Dahl RE. Understanding adolescence as a period of social-affective engagement and goal flexibility. Nat Rev Neurosci 2012;13:636–50.

69 Herting MM, Maxwell EC, Irvine C, et al. The impact of sex, puberty, and hormones on white matter microstructure in adolescents. Cereb Cortex 2012;22:1979–92.

70 Tyborowska A, Volman I, Niermann HCM, et al. Developmental shift in testosterone influence on prefrontal emotion control. Dev Sci 2024;27:e13415.

